# The immunogenicity and reactogenicity of four COVID-19 booster vaccinations against SARS-CoV-2 variants of concerns (Delta, Beta, and Omicron) following CoronaVac or ChAdOx1 nCoV-19 primary series

**DOI:** 10.1101/2021.11.29.21266947

**Authors:** Nasikarn Angkasekwinai, Suvimol Niyomnaitham, Jaturong Sewatanon, Supaporn Phumiamorn, Kasama Sukapirom, Sansnee Senawong, Zheng Quan Toh, Pinklow Umrod, Thitiporn Somporn, Supaporn Chumpol, Kanokphon Ritthitham, Yuparat Jantraphakorn, Kanjana Srisutthisamphan, Kulkanya Chokephaibulkit

**Author notes:** **Corresponding author:** Kulkanya Chokephaibulkit, MD Professor of Pediatrics, Department of Pediatrics, Faculty of Medicine Siriraj Hospital, Mahidol University, Siriraj Institute of Clinical Research (SICRES), 2 Wanglang Road, Bangkoknoi, Bangkok 10700, Thailand. Nasikarn Angkasekwinai and Suvimol Niyomnaitham equally contributed to the research work.

## Abstract

The CoronaVac (Sinovac Biotech) and ChAdOx1(Oxford-AstraZeneca) are two widely used COVID-19 vaccines. We examined the immunogenicity of four COVID-19 booster vaccine: BBIBP-CorV (Sinopharm Biotech), ChAdOx1, 30μg-BNT162b2 and 15μg-BNT162b2 (Pfizer-BioNTech), in healthy adults who received a two-dose CoronaVac or ChAdOx1 8-12 weeks earlier. Among the 352 participants (179 CoronaVac and 173 ChAdOx1 participants), 285 (81%) were female, and median age was 39(IQR: 31-47) years. 98%(175/179) and 99%(172/173) of Coronavac and ChAdOx1 participants remained seropositive at baseline. Two weeks post-booster, both 30μg- and 15μg-BNT162b2 induced the highest anti-RBD IgG concentration (BAU/mL); Coronavac-prime: 30μg-BNT162b2, 5152.2(95%CI 4491.7-5909.8); 15μg-BNT162b2, 3981.1(3397.2-4665.4); ChAdOx1, 1358.0(1141.8-1615.1); BBIBP-CorV, 154.6(92.11-259.47); ChAdOx1-prime: 30μg-BNT162b2, 2363.8(2005.6-2786.1; 15μg-BNT162b2, 1961.9(1624.6-2369.1); ChAdOx1, 246.4(199.6-304.2); BBIBP-CorV, 128.1(93.5-175.4). Similarly, both 30μg- and 15μg-BNT162b2 boosting induced the highest neutralizing antibodies (nAb) titres against all variants and highest T-cell response evaluated by interferon gamma released asssays. While all BNT162b2 or heterologous ChAdOx1-boosted participants had nAb against Omicron, these were <50% for BBIBP-CorV and 75% for homologous ChAdOx1-boosted participants. There was significant decrease in nAb (>4-fold) 16-20 weeks post booster. Heterologous boosting with BNT162b2 following CoronaVac or ChAdOx1 primary series is most immunogenic. A lower dose BNT162b2 may be considered as booster in settings with limited vaccine supply.

## Introduction

Both CoronaVac (an inactivated whole-virion SARS-CoV-2 vaccine, Sinovac Life Science) and ChAdOx1 (a chimpanzee adenovirus-vectored vaccine expressing the SARS-CoV-2 spike protein, Oxford, AstraZeneca) are safe and effective vaccines against symptomatic COVID-19 caused by the ancestral Wuhan strain, and to a lower extent against the Delta variant, and even lower efficacy against Omicron [1-7]. These two vaccines are widely used vaccines globally, particularly in low- and middle-income countries [8].

Breakthrough infections following COVID-19 vaccination, which are likely due to a combination of waning immunity and the emergence of SARS-CoV-2 variants, have led to the need for booster vaccination [9-13]. While the antibody threshold of protection has not been identified, higher antibody levels are likely to be associated with greater protection^7^. Cell mediated immune responses generated following vaccination also plays an important role in protection against SARS-CoV-2.

Several studies have demonstrated improved humoral responses with heterologous COVID-19 prime-boost vaccination, primarily on ChAdOx1 and mRNA vaccines [14-18]. However, other combinations of prime-boost COVID-19 vaccination involving inactivated COVID-19 vaccines have not been evaluated. Furthermore, the persistence of immunity following a booster (3rd) dose of COVID-19 vaccine is unknown. A recent study of reduced dosage of mRNA-1273 vaccine as a booster was found to be highly immunogenic, suggesting that a lower dosage vaccine may be equally immunogenic as a standard dosage, particularly for mRNA vaccines [19].

In this study, we examined the safety and immunogenicity of four booster vaccinations at 2 weeks and up to 16-20 weeks in healthy adults who previously received a 2-dose primary series of CoronaVac or ChAdOx1 vaccine 8-12 weeks earlier.

## Results

Among 352 participants enrolled (179 and 173 participants in CoronaVac- and ChAdOx1-prime group), 285 (81%) were female, and the median age was 39 (interquartile range, IQR: 31-47) years. The demographic of the study participants receiving different booster vaccine was shown in Table 1. The recruitment for BBIBP-CorV booster groups were stopped after 36 participants, 14 in CoronaVac-prime and 22 in ChAdOx1-prime, after the preliminary analysis found low anti-SARS-CoV-2 RBD IgG concentration.

**Table 1.** Baseline characteristics of participants.

|  | Type of booster vaccinations |  |  |  | *p*-value |
| --- | --- | --- | --- | --- | --- |
|  | BBIBP-CorV<br>n=14 | ChAdOx1<br>n=65 | 30 µg BNT162b2<br>n=50 | 15 µg BNT162b2<br>n=50 |  |
| **Corona Vac-prime (n=179)** |  |  |  |  |  |
| Age (years), median (IQR) | 31<br>(27, 41.5) | 36.6<br>(29.5, 44) | 32<br>(28, 41.8) | 40<br>(31.5, 45.3) | 0.018 |
| Female, n (%) | 12 (85.7) | 51 (78.5) | 40 (80.0) | 33 (66.0) | 0.249 |
| BMI (kg/m<sup>2</sup>), median (IQR) | 25.2<br>(21.1, 31.6) | 23.4<br>(20.9, 27.1) | 22.1<br>(19.5, 25.5) | 23.9<br>(20.9, 26.0) | 0.325 |
| **ChAdOx1-prime (n=173)** |  |  |  |  |  |
|  | BBIBP-CorV<br>n=23 | ChAdOx1<br>n=50 | 30 µg BNT162b2<br>n=50 | 15 µg BNT162b2<br>n=50 | *p*-value |
| Age (years), median (IQR) | 51<br>(42, 59) | 45.5<br>(36, 57) | 34<br>(30, 43) | 41.5<br>(34, 49.5) | 0.001 |
| Female, n (%) | 21 (91.3) | 47 (94.0) | 37 (74.0) | 44 (88.0) | 0.001 |
| BMI (kg/m<sup>2</sup>), median (IQR) | 24.8<br>(22.4, 27.6) | 23.8<br>(21.3, 26.7) | 21.4<br>(19.3, 24.7) | 23.3<br>(20.4, 26.5) | 0.001 |

### Adverse events (AEs)

Among the CoronaVac-prime groups, the overall AEs was most frequent after boosting with ChAdOx1 (98%), followed by 30μg-BNT162b2 (92%), 15μg-BNT162b2 (80%), and BBIBP-CorV (70%); whereas in ChAdOx1-prime group, the overall AEs was most frequent after boosting with 30μg-BNT162b2 (98%), followed by 15μg-BNT162b2 (88%), ChAdOx1 (72%), and BBIBP-CorV (61%) (Fig. 1, Supplementary Table 1). Systemic AEs were in the same trend as local AEs (Fig. 1 and Supplementary Table 1). All AEs were mild (grade 1) to moderate (grade 2) in severity and recovered within 2-3 days. No serious AEs was found in this study.

**Figure 1.**
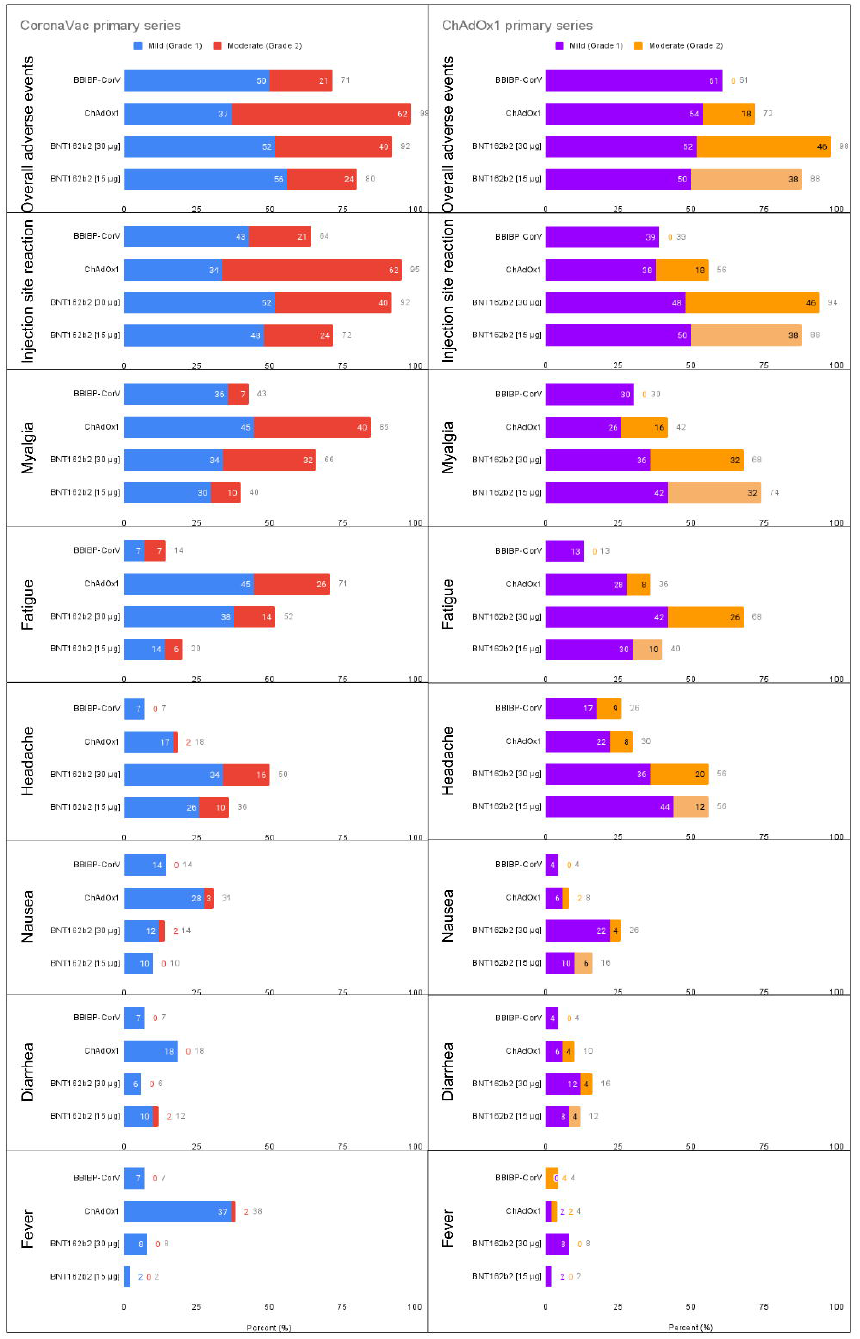
Adverse events following four different booster vaccinations. The stacked bars represent the percentage of participants who reported mild and moderate adverse events after the booster vaccinations in the subjects who had received 2-dose CoronaVac-primary series (A) and ChAdOx1-primary series (B) vaccination. Chi-square was used for statistical analyses.

### Anti-SARS-CoV-2 RBD IgG responses

At baseline (8-12 weeks post-primary series), 175/179 (97.8%) participants in CoronaVac-prime and 172/173 (99.4%) in ChAdOx1-prime remained seropositive. The anti-RBD IgG GMC at baseline were lower in the CoronaVac-prime groups than in the ChAdOx1-prime group (36.31 vs. 98.27 BAU/mL) (Fig. 2A-B). For the CoronaVac-prime groups, the anti-RBD IgG geometric mean concentrations (GMC) post-booster in the 30μg-BNT162b2 group (5152.2 BAU/mL, 95%CI 4491.7-5909.8) was significantly higher than other vaccine booster groups: 15μg-BNT162b2 (3981.1 BAU/mL, 95% CI 3397.2-4665.4), ChAdOx1 (1,358 BAU/mL, 95%CI 1141.8, 1615.1), and BBIBP-CorV (154 BAU/mL, 95%CI 92.11, 259.47) (Fig. 2A and Supplementary Table 2). The geometric mean ratio (GMR) between post-boost and post-primary series of CoronaVac for BBIBP-CorV, ChAdOx1, 30μg-BNT162b2 and 15μg-BNT162b2 were 0.94, 8.26, 31.34, and 24.22, respectively (Supplementary Table 2).

**Figure 2.**
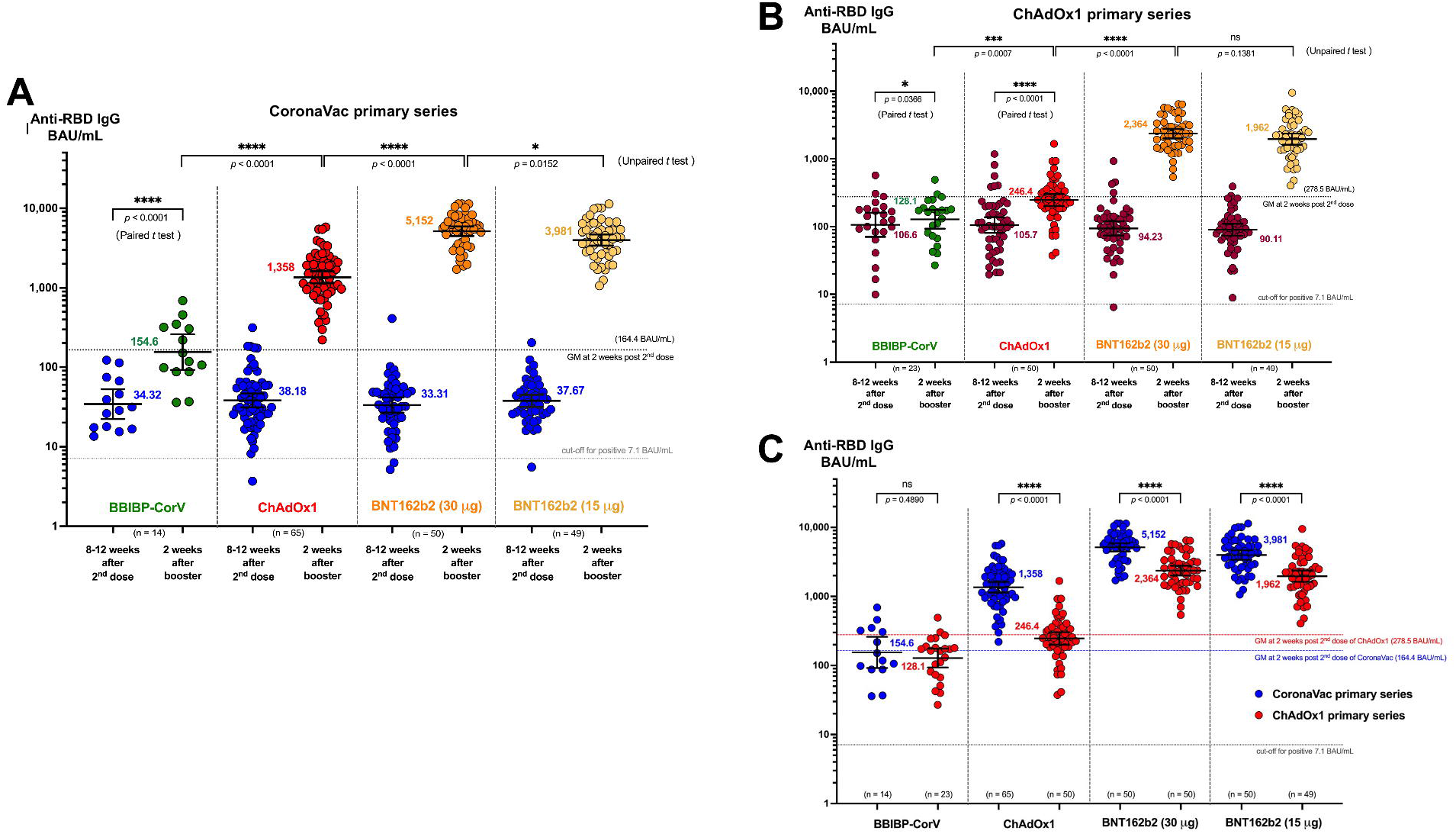
SARS-CoV-2 RBD IgG 2 weeks after booster vaccination. The scatter dot plot represents the SARS-CoV-2 RBD IgG concentration before and 2 weeks after different booster vaccination in participants who received 2-dose CoronaVac primary series (**A**) or ChAdOx1-primary series (**B**) 8-12 weeks prior. (**C**) Comparison of SARS-CoV-2 RBD IgG levels at 2 weeks after booster vaccination between participants who received CoronaVac primary series (blue) or ChAdOx1 primary series (red). Error bars represent geometric mean and 95% confidence interval. The upper dotted line represents the geometric mean concentration (GMC) of SARS-CoV-2 RBD IgG at 2 weeks after the second dose of the 2-dose primary series of CoronaVac or ChAdOx1[7]. The lower dotted line represents the cut-off level for seropositivity.

For the ChAdOx1-prime group, the anti-RBD IgG GMC post-booster was significantly higher in participants who received 30μg-BNT162b2 (2363.8, 95%CI 2005.6-2786.1) or 15μg-BNT162b2 (1961.9 BAU/mL, 95% CI 1624.6-2369.1) compared to those who received ChAdOx1 (246.4 BAU/mL, 95%CI 199.6-304.2); and BBIBP-CorV (128.1 BAU/mL, 95%CI 93.5-175.4) (Fig. 2B). The GMR between post-boost and post-primary series of ChAdOx1 were 0.46, 0.88, 8.49, and 7.04 for BBIBP-CorV, ChAdOx1, 30μg-BNT162b2 and 15μg-BNT162b2, respectively (Supplementary Table 2). The post-boost GMC levels in ChAdOx1-prime were generally lower than that in the CoronaVac-prime group for all booster vaccines (Fig. 2C).

### Neutralizing antibody responses against the SARS-CoV-2 variants

At 2 weeks post booster dose, almost all participants had (50% plaque reduction neutralization titre) PRNT_50_ against Delta and Beta; 1/30 (3%) participant in the ChAdOx1-ChAdOx1 group was negative against Delta and nine participants among the CoronaVac-BBIBP-CorV (2/14, 14%), ChAdOx1-BBIBP-CorV (3/22, 14%) and ChAdOx1-ChAdOx1 (4/30, 13%) were negative against Beta. For both the CoronaVac-prime and ChAdOx1-prime groups, the PRNT_50_ GMT against the Delta (Fig. 3A) and Beta (Fig. 3B) variant were significantly higher among those who received a booster dose of BNT162b2 (30μg or 15μg) compared to those who received ChAdOx1 or BBIBP-CorV. There was no statistical difference in PRNT_50_ between boosting with 30μg and 15μg-BNT162b2 regardless of the primary series vaccine and the type of variants. However, the PRNT_50_ against the Beta variant was in general around 1.5-fold lower than the Delta variants for both CoronaVac-prime and ChAdOx1-prime groups (Fig. 3C). The GMRs of the PRNT_50_ between post-boost and post-primary series were highest among the participants who received BNT162b2 boosting vaccination in both CoronaVac-prime and ChAdOx1-prime groups (Table 2). The SARS-CoV-2 RBD IgG levels and the PRNT_50_ against Delta variant (Supplementary Fig. S1A and B) or Beta variant (Supplementary Fig. S1C and D) were strongly correlated (r = 0.49-0.89).

**Table 2.**
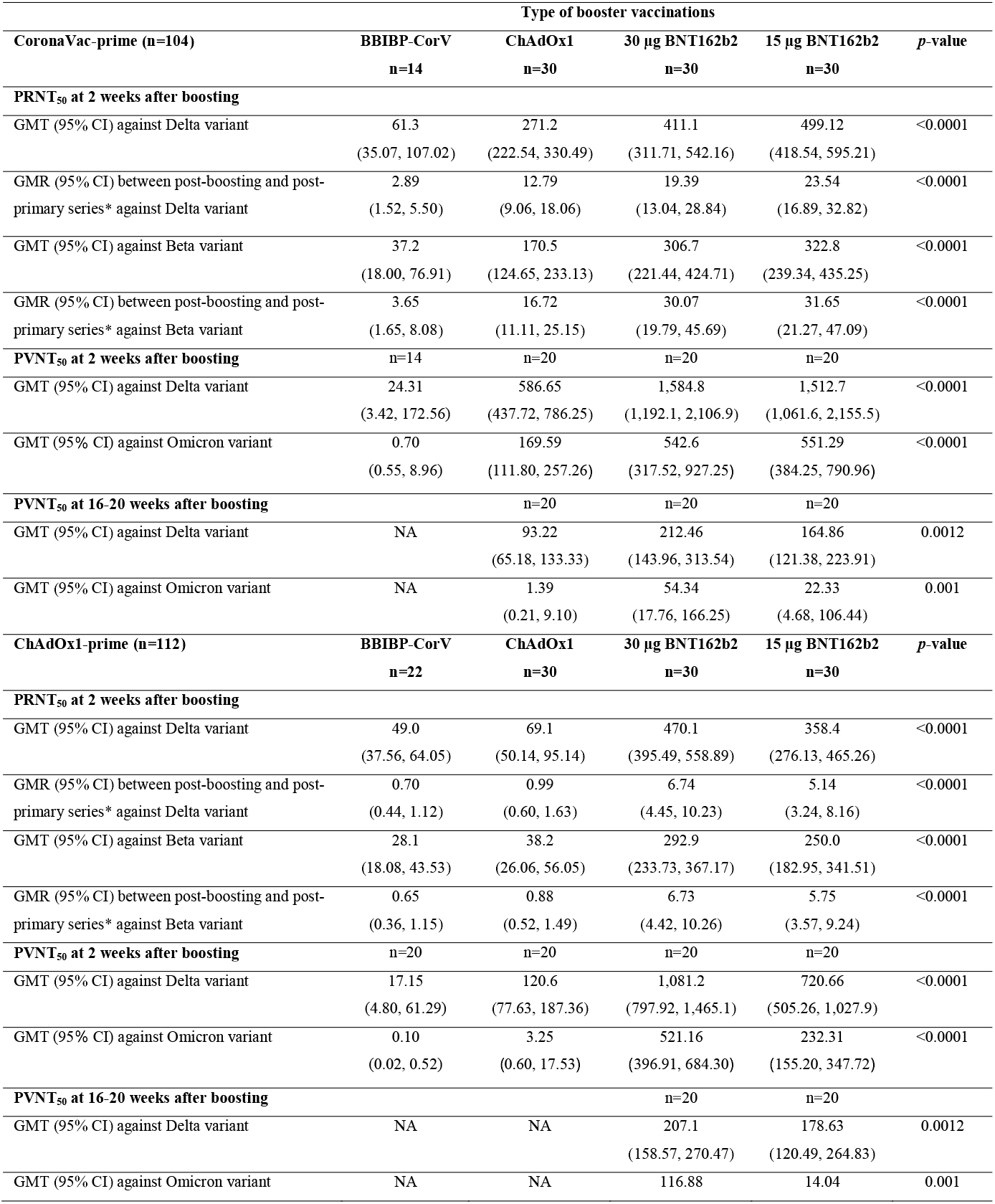
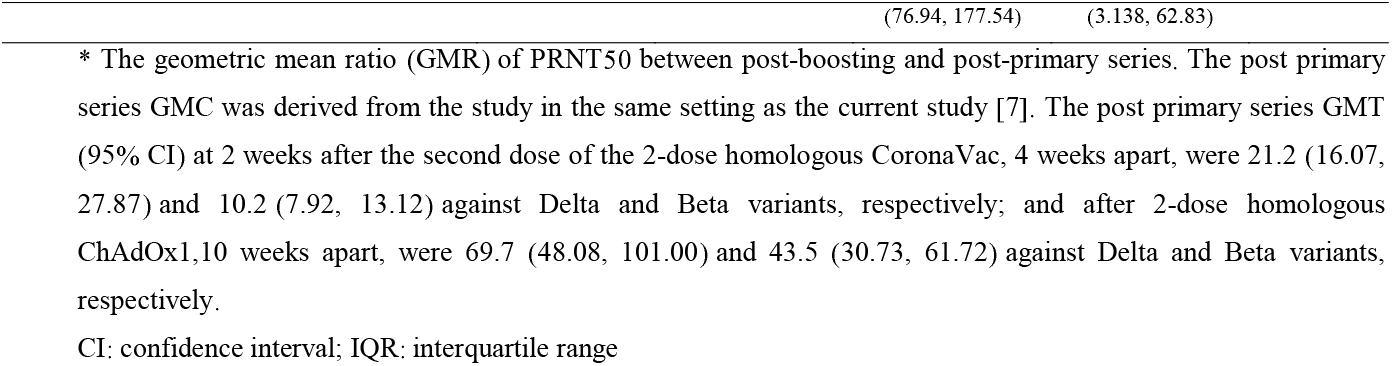
The 50% plaque reduction neutralization (PRNT_50_) and 50% pseudovirus neutralization (PVNT_50_) geometric mean antibody titers (GMT) against variant.

**Figure 3.**
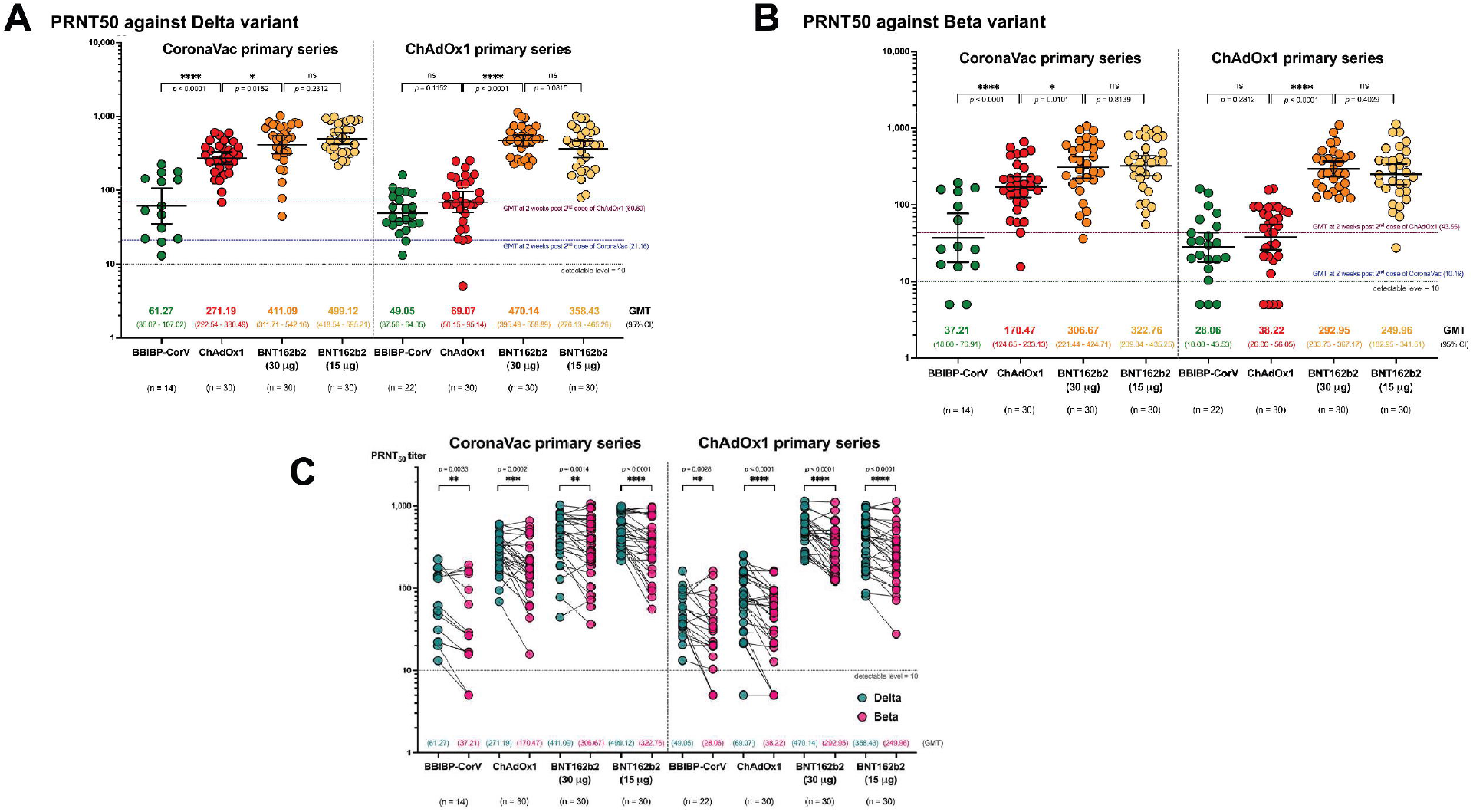
Plaque reduction neutralization titers (PRNT_50_) for SARS-CoV-2 Delta and Beta variants. Scatter dot plots represent PRNT_50_ titer against the (**A**) Delta or (**B**) Beta variant at 2 weeks after different booster vaccines in participants who received two doses of Coronavac or ChAdOx1 8-12 weeks earlier. (**C**) Comparison of PRNT_50_ between SARS-CoV-2 Delta (green) and Beta (pink) variants 2 weeks after booster vaccination. Error bars represent geometric mean titer (GMT) and 95% confidence interval (CI). The upper dotted line represents the geometric mean values of anti-SARS-CoV-2 RBD IgG at 2 weeks after the second dose of the 2-dose primary series of CoronaVac or ChAdOx1 [7]. Lower dot line represents the cut-off level for seropositivity.

In order to compare the neutralising titers between Delta and Omicron, we performed the pseudovirion neutralization test (PVNT) assay on both variants. At 2 weeks post booster dose, almost all participants had 50% pseudovirus neutralization antibody titres (PRNT_50_) against Delta, except for 4 participants in the CoronaVac-BBIBP-CorV (2/14, 14%) and ChAdOx1-BBIBP-CorV (2/20, 10%). In contrast, PRNT_50_ against Omicron was only present in ≤50% in CoronaVac-BBIBP-CorV and ChAdOx1-BBIBP-CorV groups, and 75% (15/20) in the ChAdOx1-ChAdOx1. Among the CoronaVac-prime groups, 15μg and 30μg-BNT162b2 booster induced similar PVNT_50_ against Omicron (Fig 4A), whereas in the ChAdOx1-prime groups, the group that received 15μg-BNT162b2 induced significantly lower PVNT_50_ against Omicron compared to the 30μg-BNT162b2 group (Fig. 4B). Notably, both CoronaVac- and ChAdOx1-prime groups that received ChAdOx1 booster had significantly lower PVNT_50_ against Delta and Omicron variants than the groups that received 15μg-or 30 μg BNT162b2 (Fig. 4A and 4B). Between the CoronaVac- and ChAdOx1-prime groups that received ChAdOx1 booster, CoronaVac prime-ChAdOx1 boost group had significantly higher PVNT_50_ against Delta and Omicron variants than the ChAdOx1 prime-ChAdOx1 boost group (Fig. 4A-B and Table 2). The PVNT_50_ GMT against Omicron was 2-to 37-folds lower than that against Delta (Fig. 4C and Table 2).

**Figure 4.**
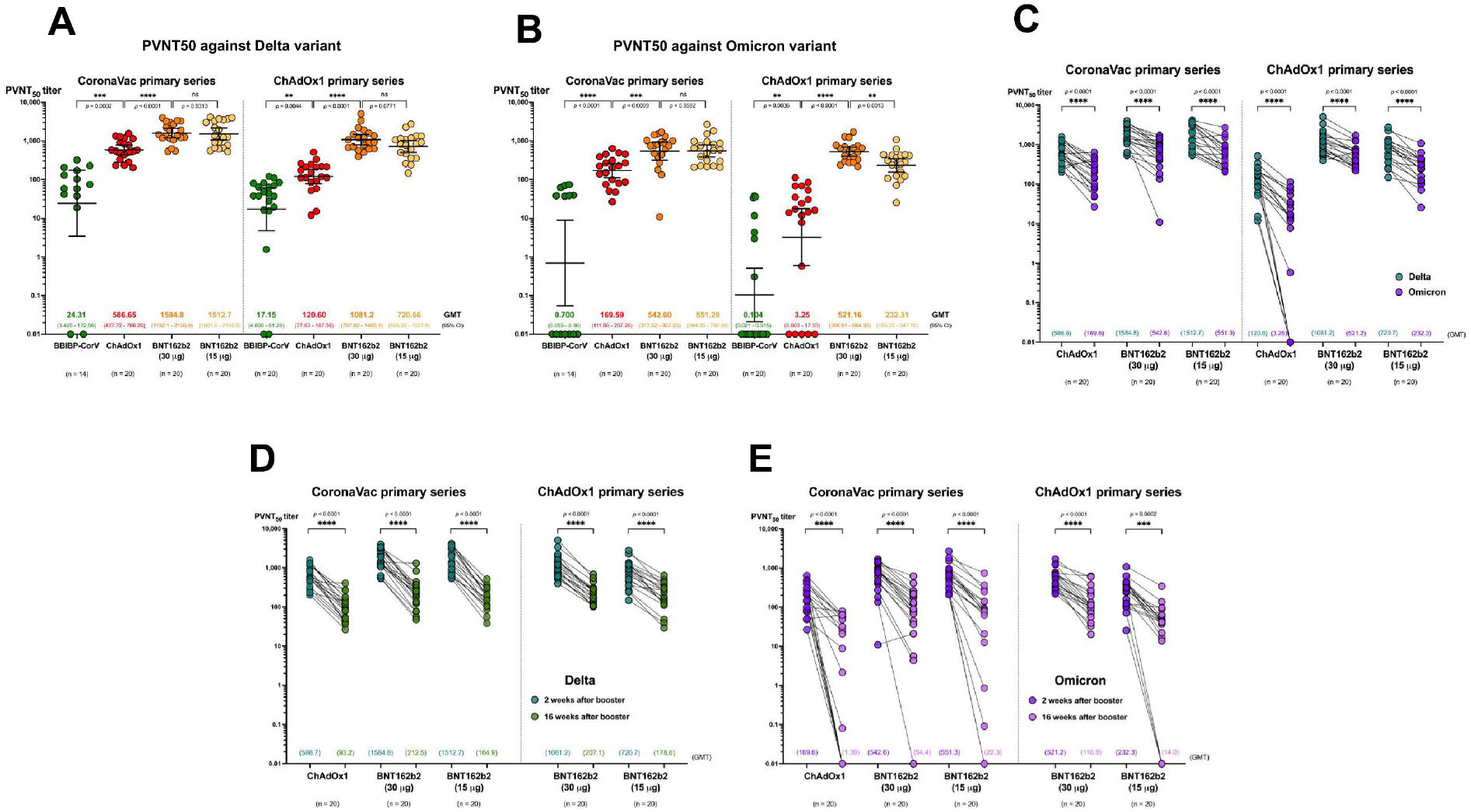
Pseudovirion neutralization titers (PVNT_50_) for SARS-CoV-2 Delta and Omicron variants. Aligned dot plots represent PVNT_50_ against the (**A**) Delta or (**B**) Omicron variant at 2 weeks after different booster vaccines in participants who received two doses of Coronavac or ChAdOx1 8-12 weeks earlier. (**C**) Comparison of PRNT_50_ between SARS-CoV-2 Delta (green) and Omicron (purple) variants 2 weeks after booster vaccination. PVNT_50_ titer against the Delta **(D)** or (**E**) Omicron variant at 2 weeks and 16-20 weeks of the same participants after different booster vaccines. Error bars represent geometric mean titer (GMT) and 95% confidence interval (CI).

The groups that received BBIPB-CorV as booster or ChAdOx1 as priming and booster (3-dose ChAdOx1) were not followed up for the 16-20 weeks as they have received additional booster vaccination outside of this study after revealing the low antibody results. For the rest of the groups, there was a significant decline (at least 4-fold) in PVNT_50_ against Delta and Omicron at 16-20 weeks after boosting in both the CoronaVac-prime and ChAdOx1-prime groups (Fig. 4D-E and Table 2). However, 100% and >90% of each group remained seropositive against Delta and Omicron. No significant difference in PVNT_50_ against Delta and Omicron was observed between the CoronaVac-prime and ChAdOx1-prime groups at this timepoint (Table 2). However, a more significant drop in PVNT_50_ against Omicron (4.5 to 122 folds) was observed compared to the Delta variant (4 to 9-fold) (Fig 4D-E, Table 2).

### QuantiFERON SARS-CoV-2 interferon gamma release assay (IGRA)

Cellular immunity was measured at baseline using the QuantiFERON SARS-CoV-2 interferon gamma release assay (IGRA). Participants with a negative IGRA response at baseline were tested again at two weeks post-booster. At baseline, a higher proportion of 35.8% (62/173) of participants in ChAdOx1-prime group and 25% (45/179) of CoronaVac-prime group had positive IGRA (*P*=0.029). Among those with negative IGRA at baseline, IGRA conversion was the highest after a booster dose of 30μg-BNT162b2, followed by 15μg-BNT162b2, ChAdOx1, and BBIBP-CorV (Supplementary Table 2). None of the study participants who were IGRA-negative at baseline in the ChAdOx1-prime group had a positive IGRA response following boosting with BBIBP-CorV or ChAdOx1 (Supplementary Table 2 and Supplementary Fig. S2).

## Discussion

In this study, BBIBP-CorV, ChAdOx1, BNT162b2 (standard and reduced dosage) given as booster dose to individuals who previously received either CoronaVac or ChAdOx1 primary series were found to be safe and well tolerated. BNT162b2 given as a booster induced the highest humoral and cellular immune responses compared to BBIBP-CorV or ChAdOx1. Furthermore, both 15μg and 30μg-BNT162b2 induced similar humoral responses against the SARS-CoV-2 all variants tested for both CoronaVac- and ChAdOx1-prime groups, except for the neutralising antibody titers against the Omicron variant in the ChAdOx1-prime group. Notably, higher humoral response was observed in the CoronaVac-prime group following the booster dose compared to the ChAdOx1-prime group while having the lower circulating antibodies at baseline. Despite a rapid decline in neutralising antibodies against Delta and Omicron 16-20 weeks following heterologous ChAdOx1 or BNT162b2 booster, a high proportion of individuals still have antibodies against Delta and Omicron.

Heterologous boosting vaccination in our study were generally well tolerated, and the AEs rates observed in this study were in line with those reported in COVID-19 vaccine primary series and booster studies [19,20]. Heterologous boosting regimen were also found to be more immunogenic than homologous ChAdOx1 boosting regimen or homologous inactivated vaccines regimen (CoronaVac prime-BBIBP-CorV boost) in our study, which was consistent with recent studies [21-25]. However, heterologous boosting with BBIBP-CorV vaccine was poorly immunogenic, which was in line with previous studies, including a study that revealed poor immunogenicity of heterologous ChAdOx1 prime-VLA2001 (inactivated vaccine by Valvena) boost [24]. Our findings suggest that inactivated whole virus vaccine as a booster vaccine may not be effective at generating high levels of neutralising antibodies.

The SARS-CoV-2 Omicron variant, recently identified in November 2021 has been reported to evade immunity induced from past infection or two vaccine doses [26-28]. Our results suggest that a third dose of BNT162b2 can overcome this immune evasion through the induction of neutralising antibodies. A recent study also reported high antibodies against Omicron following a third dose of mRNA vaccines (mRNA-1273 or BNT162b2) [29]. Heterologous boost with ChAdOx1 was immunogenic in CoronaVac-prime participants but was poorly immunogenic in ChAdOx1-prime recipeints. Taken together, these data support the use of BNT162b2 as a booster regardless of the primary series against the Delta and Omicron variants that are widely circulating globally. Consequently, ChAdOx1 may also be use as a booster for CoronaVac-prime participants.

The persistence of immunity following COVID-19 booster is unknown. Our findings suggest possible protection against Delta and Omicron infection for at least 16-20 weeks despite rapid waning antibody levels. It is important to note that the antibody threshold of protection against infection and severe disease has not been identified, and immune memory cells which are thought to be important for long-term protection was not measured in our study. Furthermore, a recent study reported breakthrough infections two months after receiving a mRNA booster dose (received mRNA primar series) [30]. Larger studies with longer duration are needed to confirm our findings and also determine the persistence of immunity against SARS-CoV-2 infection and severe disease. A fourth booster dose study has been studied in high-risk groups [31] and is currently under investigation in Israel [32].

Virus-specific memory T cells are important for protection against SARS-CoV-2, particularly against severe disease. Only a third of ChAdOx1-prime and a quarter of CoronaVac-prime participants in our study remained positive for IGRA as a marker for T cell response at baseline; i.e. 8-12 weeks post primary series. Previous studies evaluating 2-dose ChAdOx1 primary series have reported the generation of robust T cell response following the first dose, with no significant increase in T cell responses following the second dose [33,34], and following a homologous ChAdOx1 booster [21]. On the other hand, the study of 2-dose CoronaVac primary series revealed poor inducer of T-cell response [35]. The discrepancy in T cell responses after primary series from our study could be due to waning immunity, population differences and the different assays used to measure IFN-LJ response (Quantiferon vs. IFN-LJ ELISPOT). We found BBIBP-CorV boosting poorly induced IGRA response; however, it is important to note that inactivated vaccine (i.e. BBIBP-CorV) may have other antigens (i.e., M or N proteins) that induce T cell responses [36], whereas in our study, we only examined T cell responses to S protein, and thus may have underestimated the cellular responses. The low T cell boosting responses following homologous boosting regimen of ChAdOx1 is in line with the low neutralizing antibody boosting responses observed in this study. This could be explained by the anti-vector interference, and possibly due to a short interval (8-12 weeks) between the third and second dose.

Our finding that half-dose BNT162b2 was equally immunogenic as the standard dosage, but with less reactogenicity, suggesting that less amount of antigen may be sufficient for boosting immune responses against SARS-CoV-2. This finding is in concordance with previous study on mRNA1273 vaccine where half dose of the mRNA1273 (50 μg) was able to induce significantly higher neutralizing antibodies than the level induced after primary series against the SARS-CoV-2 variants of concerns [19]. A lower mRNA vaccine dose may be considered for COVID-19 booster vaccination, given that the limited vaccine supply globally.

There are some limitations in this study. First, our study was conducted in a non-randomized open label manner which was due to the availability of each vaccine at a different timing may lead to selection bias. Second, our sample size is small, particularly those who received BBIBP-CorV as booster; therefore, the data need to interpret with caution. Third, the participants in this study were healthy adults, and may not be generalized to other populations such as immunocompromised individuals. Lastly, how our findings translate to disease protection warrant further investigation.

In conclusion, our study found that a booster dose of BNT162b2 given to individuals previously vaccinated with CoronaVac or ChAdOx1 is the most immunogenic and induced high cross protective antibodies against Delta, Beta, and Omicron variants, and T-cell response. BBIBP-CorV and homologous ChAdOx1 are not effective booster vaccines. The rapid decline of antibodies after 16-20 weeks of receiving the booster warrants further investigation into the efficacy and persistence of immunity following the booster dose. Our study findings have important implications on the choice of booster dose for countries that have introduced CoronaVac or ChAdOx1 as primary series to date. Our study also suggests that reduced dosage of BNT162b2 may be used as a booster dose that may be highly relevant for countries with limited vaccine supply particularly if CoronaVac was used in the primary series.

## Methods

### Study design and participants

This single-center prospective, non-randomized, open-labeled cohort study enrolled 352 healthy adults, aged 18 years or older at Siriraj Hospital, a university-based referral center in Bangkok, Thailand, from July to September 2021. The eligible participants were those who have received either 2 doses of CoronaVac (4 weeks apart) (CoronaVac-prime) or ChAdOx1 (8-10 weeks apart) (ChAdOx1-prime) primary series vaccination 8-12 weeks prior to recruitment. The exclusion criteria were history of SARS-CoV-2 infection; prior received prophylactic or investigational treatment against COVID-19 within 90 days; had an unstable underlying disease; history of vaccine anaphylaxis; being pregnant; immunocompromised or currently receiving immunosuppressive agents. Written informed consent was obtained from all study participants. The study protocol was approved by the Siriraj Institutional Review Board (COA no. Si 537/2021). The study was registered in thaichinicaltrials.org (TCTR20210719006).

### Study Procedures

Eligible participants were openly assigned to receive one of the four intramuscular booster vaccinations: BBIBP-CorV (Sinopharm), ChAdOx1 (AstraZeneca), full dose [30 μg] or half dose [15 μg] BNT162b2 (Pfizer). Due to the shortage of study vaccines during the peak of the outbreak when the enrollment started, the study vaccine was assigned to the participant by order of confirmation to participate in the study and the type of vaccine available on that day.

After about 4 weeks of enrollment, the BBIBP-CorV booster group was terminated after the preliminary analysis that found low anti-SARS-CoV-2 RBD concentration.

The participants were observed for at least 30 min following vaccination for any immediate adverse events (AE) and were instructed to record self-assessment signs or symptoms in an electronic diary (eDiary) for seven days after vaccination. An AE were defined as described in the previous study [7].

Blood samples were collected at baseline (pre-booster), two weeks, and 16-20 weeks after booster vaccination to determine the anti-SARS-CoV-2 RBD IgG antibody levels. A subset of samples at two weeks and 16-20 weeks post-booster were tested for neutralizing antibodies against the SARS-CoV-2 Delta and Beta variants using the 50% plaque reduction neutralization test (PRNT_50_) and against Delta and Omicron variants using the pseudovirus neutralization test (PVNT). The groups that received BBIPB-CorV as booster or ChAdOx1 as priming and booster (3-dose ChAdOx1) were not followed up for the 16-20 weeks analysis; the participants have received additional booster vaccination outside of this study at approximately 4 weeks after receiving the study vaccination due to the low antibody response. Cellular immunity was measured at baseline using the QuantiFERON SARS-CoV-2 interferon gamma release assay (IGRA). Participants with a negative IGRA response at baseline were tested again at two weeks post-booster.

### Laboratory Assays

#### Chemiluminescent microparticle assay (CMIA) for anti-SARS-CoV-2 RBD IgG

The anti-RBD IgG was measured by CMIA using the SARS-CoV-2 IgG II Quant (Abbott, List No. 06S60) on the ARCHITECT I System as described in previous study^7^. Samples with a value >11,360 BAU/mL were reported as 11,360 BAU/mL.

#### 50% plaque reduction neutralization test (PRNT)

The standard live virus 50% plaque reduction neutralization test (PRNT_50_) against Delta variant (B.1.617.2) and Beta variant (B.1.351) were performed as described in the previous study^7^. The PRNT_50_ titer is defined as the the highest test serum dilution for which the virus infectivity is reduced by 50% when compared with the average plaque counts of the virus control (no serum). The PRNT_50_ titer of 5 was used for all samples that were below the detectable level (1:10).

#### Pseudovirus neutralization assay (PVNT)

Codon-optimized gene encoding the spike of Omicron (B.1.1.529/ BA.1) and Delta (B.1.617.2) were generated by gene synthesis (Genscript) and cloned into the pCAGGS expressing plasmid by In-Fusion assembly (Clontech). Pseudovirus was generated and concentrated as previously described [37]. Pseudotype-based neutralization assays were carried out as described previously [37]. The 50% pseudovirion neutralizing antibody titer (PVNT_50_) was calculated by interpolating the point at which infectivity was reduced to 50% of the value for the control samples (no serum).

#### QuantiFERON SARS-CoV-2 interferon gamma release assay (IGRA)

SARS-CoV-2 specific T cell responses were assessed by whole blood IGRA using QIAGEN’s proprietary mixes of SARS-CoV-2 S protein designed for CD4+ T cell (Ag1), CD8+ T cells (Ag2) according to the manufacturer’s instruction. Interferon-gamma (IFN-LJ) concentration was measured with an automated QuantiFERON SARS-CoV-2 ELISA instrument and reported in International Units per mL (IU/mL) [38,39]. The cut-off for positivity was determined as the level above the mean plus three standard deviations of the negative control. The cut-offs for Ag1 (>0.12 IU/mL) and Ag2 (>0.17 IU/mL) were determined based on 61 SARS-CoV-2 negative control samples. A positive response to either of the two peptides pools was considered positive.

### Statistical Analysis

The sample size was calculated using the lower bounds of anti-RBD IgG geometric mean concentration (GMC) from previous study[7]. A sample size of 50 participants in each group would provide us 80% power to detect any difference between groups.

The AEs endpoints were presented as frequencies and Chi-square test was used to test for statistical difference. The anti-SARS-CoV-2 RBD IgG concentration and neutralization antibodies were reported as GMC and geometric mean titers (GMT) with 95% confidence interval (CI), respectively. Anti-RBD IgG GMC and PRNT_50_ GMTs at two weeks after the primary series (post-primary series) from our previous study was used for comparison [7]: the anti-RBD IgG GMC for CoronaVac and ChAdOx1 was 164.4 BAU/mL and 278.5 BAU/mL, respecitively and the PRNT_50_ GMT was 21.2 and 69.7 for Delta variant and 10.2 and 43.5 for Beta variant, respectively [7]. The geometric mean ratio (GMR) with 95% CI was analyzed between the post-boosting levels or titers and post-primary series levels or titers references. Paired *t* test, unpaired *t* test, and analysis of variance (ANOVA) were used to compare GMC and GMT within group, between groups, and across groups using GraphPad Prism 9 version 9.2.0 (283) (GraphPad Software, CA, USA), respectively. Other statistical analyses were conducted using STATA version 17 (Stata Corp, LP, College Station, TX, USA).

## Data Availability

The datasets generated during and/or analyzed during the current study are available from the corresponding author on reasonable request.

## Acknowledgement

The authors gratefully acknowledge the Siriraj Institute of Clinical Research (SICRES) team, Abbott Laboratories Ltd. for technical supports and all health care workers who took part and enabled this study to be possible. We are also grateful to Professors Kim Mulholland and Paul Licciardi, Murdoch Children’s Research Institute, who provided review and critical comments to improve the study and the manuscript.

## Author Contributions

N.A. and S.N. equally contributed to the research work. Conceptualization and Methodology: N.A., S.N., J.S., K.R., Y.J, K.S; Formal analysis and data curation: N.A., J.S., S.N. Z.Q.T.; Project administration, N.A, J.S., S.N.; Supervision, K.C.; Resources and Funding, K.C. All authors involved with investigation, and writing-review and editing.

## Data availability statement

Data are available upon reasonable request.

---

**Supplementary Figure S1.**
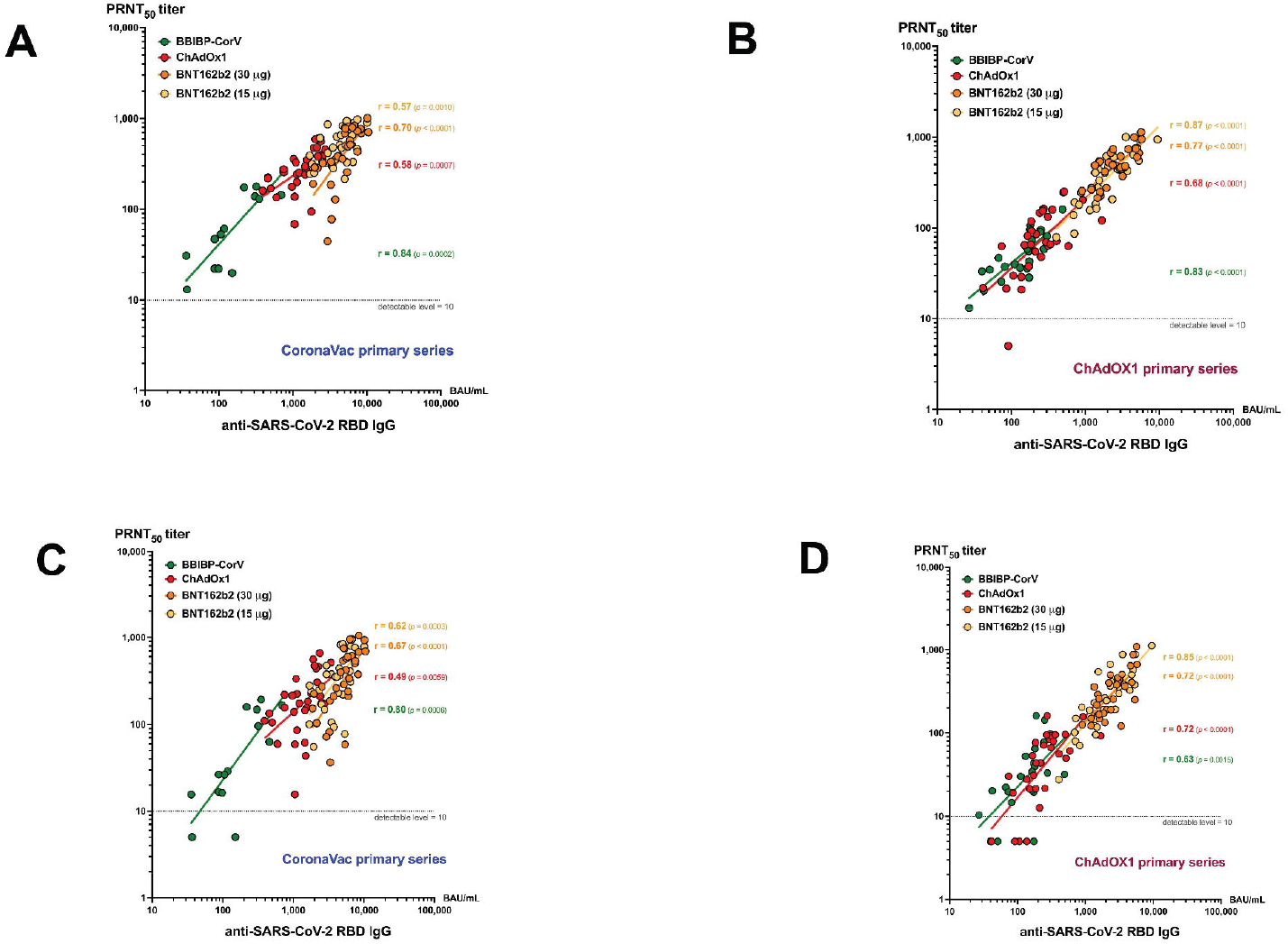
Correlation between the level of anti-SARS-CoV-2 RBD IgG and plaque reduction neutralization test (PRNT_50_) titers for the SARS-CoV-2 Delta and Beta variants. Dot plots show the correlation between the level of anti-SARS-CoV-2 RBD IgG and PRNT_50_ titer against the Delta participants who have previously received two doses of CoronaVac (A) or ChAdOx1 (B) or Beta variant in participants who had completed two doses of CoronaVac (C) or ChAdOx1 (D) 2 weeks after booster with BBIBP-CorV (green), ChAdOX1 (red), 30 μg BNT162b2 (orange) and 15 μg BNT162b2 (yellow). Pearson’s correlation coefficient (r) with *p* value for each booster vaccine indicated.

**Supplementary Figure S2.**
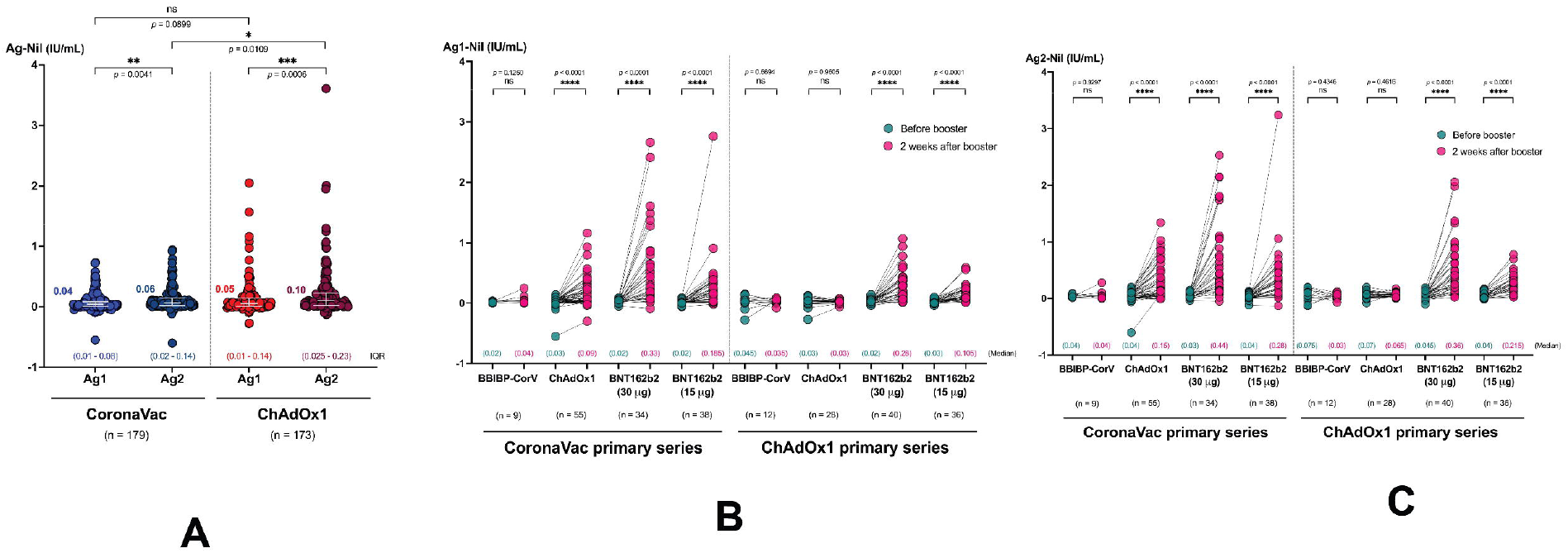
Cellular immune responses by interferon-gamma (IFN*γ*) releasing assay (IGRA). (A) Scatter dot plots represent the level of IFN*γ* following stimulation with either Ag1 or Ag2 at 8-12 weeks after two doses of CoronaVac or ChAdOx1 (before booster vaccination). Aligned dot plots show the level of IFNY following stimulation with stimulated with either (**B**) Ag1 or (**C**) Ag2 in samples collected before (teal) and 2 weeks after (pink) booster with BBIBP-CorV, ChAdOX1, 30 μg BNT162b2 and 15 μg BNT162b2. Median and interquartile range (IQR) of each group are indicated. IU/mL: international units per mL.

**Supplementary Table 1.** Adverse events of following booster vaccination.

| Adverse events (AEs) | BBIBP-CorV | ChAdOx1 | 30 µg<br>BNT162b2 | 15 µg<br>BNT162b2 | *p*-value |
| --- | --- | --- | --- | --- | --- |
| **CoronaVac-prime (n=179)** | **n=14** | **n=65** | **n=50** | **n=50** |  |
| Overall AEs (%) | 10 (71.43) | 64 (98.46) | 46 (92.0) | 40 (80.0) | 0.002 |
| Injection site reaction (%) | 9 (64.29) | 62 (95.38) | 46 (92.00) | 36 (72.00) | <0.001 |
| Fatigue (%) | 2 (14.29) | 46 (70.77) | 26 (52.0) | 10 (20.00) | <0.001 |
| Headache (%) | 1 (7.14) | 12 (18.46) | 25 (50.0) | 18 (36.0) | <0.001 |
| Myalgia (%) | 6 (42.86) | 54 (83.08) | 2 (4.0) | 1 (2.0) | <0.001 |
| Malaise (%) | 0 | 1 (1.54) | 31 (62.0) | 20 (40.0) | <0.001 |
| Nausea (%) | 2 (14.29) | 20 (30.77) | 7 (14.0) | 5 (10.0) | 0.119 |
| Diarrhea (%) | 1 (7.14) | 12 (18.46) | 3 (6.0) | 6 (12.0) | 0.284 |
| Fever (%) | 1 (7.14) | 25 (38.46) | 4 (8.0) | 1 (2.0) | <0.001 |
| Rash (%) | 2 (14.29) | 9 (13.85) | 7 (14.0) | 2 (4.0) | 0.341 |
| Somnolence (%) | 0 | 0 | 2 (4.0) | 2 (4.0) | 0.357 |
| Flu-like symptoms (%) | 0 | 4 (6.15) | 1 (2.0) | 1 (2.0) | 0.453 |
| Arthralgia (%) | 0 | 2 (3.08) | 1 (2.0) | 1 (2.0) | 0.906 |
| Dizziness (%) | 0 | 1 (1.54) | 1 (2.0) | 0 | 0.758 |
| Paresthesia (%) | 2 (14.29) | 1 (1.54) | 1 (2.0) | 0 | 0.014 |
| Vomiting (%) | 0 | 4 (6.15) | 0 | 1 (2.0) | 0.199 |
| **ChAdOx1-prime (n=173)** | **n=23** | **n=50** | **n=50** | **n=50** |  |
| Overall AEs (%) | 14 (60.87) | 36 (72.0) | 49 (958.0) | 44 (89.0) | <0.001 |
| Injection site reaction (%) | 9 (39.13) | 28 (56.0) | 47 (94.00) | 44 (88.00) | <0.001 |
| Fatigue (%) | 3 (13.04) | 18 (36.0) | 34 (68.0) | 20 (40.00) | <0.001 |
| Headache (%) | 6 (26.09) | 15 (30.0) | 28 (56.0) | 28 (56.0) | 0.024 |
| Myalgia (%) | 6 (42.86) | 54 (83.08) | 2 (4.0) | 1 (2.0) | <0.001 |
| Malaise (%) | 7 (30.43) | 21 (42.0) | 34 (68.0) | 37 (74) | 0.001 |
| Nausea (%) | 1 (4.35) | 20 (30.77) | 7 (14.0) | 5 (10.0) | 0.102 |
| Diarrhea (%) | 1 (4.35) | 5 (10.0) | 8 (16.0) | 6 (10.0) | 0.834 |
| Fever (%) | 1 (4.35) | 2 (4.0) | 4 (8.0) | 1 (2.0) | 0.240 |
| Rash (%) | 0 | 1 (2.0) | 1 (2.0) | 0 | 0.688 |
| Somnolence (%) | 0 | 1 (2.0) | 2 (4.0) | 0 | 0.421 |
| Flu like symptoms (%) | 0 | 1 (2.0) | 2 (4.0) | 2 (4.0) | 0.738 |
| Arthralgia (%) | 0 | 0 | 0 | 1 (2.0) | 0.480 |
| Dizziness (%) | 2 (8.70) | 1 (2.0) | 0 | 0 | 0.118 |
| Paresthesia (%) | 0 | 0 | 0 | 0 | - |
| Vomiting (%) | 0 | 3 (6.0) | 0 | 0 | 0.057 |

**Supplementary Table 2.** Anti-RBD IgG geometric mean concentration (GMC) and the geometric mean ratio (GMR) between post boosting and pre-boosting (baseline) or post primary series* and IGRA positive rate.

| Anti-RBD IgG geometric mean concentration<br>(GMC), BAU/mL | Type of booster vaccinations |  |  |  |  |
| --- | --- | --- | --- | --- | --- |
| CoronaVac-prime (n=179) |  |  |  |  |  |
|  | BBIBP-CorV<br>n=14 | ChAdOx1<br>n=65 | 30 µg BNT162b2<br>n=50 | 15 µg BNT162b2<br>n=50 | *p*-value<br>between<br>groups |
| GMC (95%CI) at baseline | 34.32<br>(22.33, 52.77) | 38.18<br>(31.21, 46.71) | 33.31<br>(26.72, 41.53) | 37.67<br>(31.65, 44.84) | 0.7616 |
| GMC (95%CI) at 2 weeks after boosting | 154.6<br>(92.11, 259.47) | 1358.0<br>(1141.84, 1615.07) | 5152.2<br>(4491.65, 5909.83) | 3981.1<br>(3397.15, 4665.42) | <0.0001 |
| GMR (95% CI) between 2 weeks after boosting<br>and baseline | 4.5<br>(2.98, 6.80) | 35.6<br>(29.18, 43.34) | 154.7<br>(124.30, 192.50) | 105.7<br>(90.31, 123.68) | <0.0001 |
| GMR (95% CI) between 2 weeks after boosting<br>and 2 weeks after primary series of CoronaVac* | 0.94<br>(0.53, 1.67) | 8.26<br>(6.29, 10.85) | 31.34<br>(24.37, 40.30) | 24.22<br>(18.60, 31.54) | <0.0001 |
| GMC (95%CI) at 16-20 weeks after boosting | NA | 291.32<br>(247.92, 342.33) | 774.85<br>(653.33, 918.98) | 525.31<br>(428.37, 644.18) | <0.0001 |
| Baseline SARS-CoV-2 IGRA positive, n (%) | 5 (35.7) | 12 (18.5) | 16 (32) | 12 (24) | 0.301 |
| Post-boosting IGRA positive among baseline<br>negative participants, n (%) | 1/9<br>(11.1) | 26/53<br>(49.1) | 28/34<br>(82.4) | 30/38<br>(79.0) | <0.0001 |
| ChAdOx1-prime (n=173) |  |  |  |  |  |
|  | BBIBP-CorV<br>n=23 | ChAdOx1<br>n=50 | 30 µg BNT162b2<br>n=49 | 15 µg BNT162b2<br>n=50 | *p*-value<br>between<br>groups |
| GMC (95%CI) at baseline | 106.6<br>(70.89, 160.29) | 105.7<br>(80.97, 137.97) | 95.98<br>(75.84, 121.45) | 90.11<br>(73.62, 110.30) | 0.7661 |
| GMC (95%CI) 2 weeks after boosting | 128.1<br>(93.52, 175.37) | 246.4<br>(199.59, 304.20) | 2363.8<br>(2005.58, 2786.06) | 1961.9<br>(1624.61, 2369.10) | <0.0001 |
| GMR (95% CI) between 2 weeks after boosting<br>and baseline | 1.2<br>(1.01, 1.43) | 2.3<br>(1.92, 2.83) | 25.1<br>(20.30, 31.01) | 21.8<br>(18.28, 25.92) | <0.0001 |
| GMR (95% CI) between 2 weeks after boosting<br>and 2 weeks after primary series of ChAdOx1* | 0.46<br>(0.28, 0.65) | 0.88<br>(0.58, 1.13) | 8.49<br>(5.71, 10.44) | 7.04<br>(4.69, 8.84) | <0.0001 |
| GMC (95%CI) at 16-20 weeks after boosting | NA | NA | 431.11<br>(367.59, 505.60) | 314.43<br>(267.01, 370.27) | 0.0066 |
| Baseline SARS-CoV-2 IGRA positive, n (%) | 13 (56.5) | 26 (52.0) | 9 (18.0) | 14 (28.0) | <0.0001 |
| Post-boosting IGRA positive among baseline<br>negative participants, n (%) | 0/10<br>(0) | 0/24<br>(0) | 31/41<br>(75.6) | 24/26<br>(66.7) | <0.0001 |

\*The post primary series GMC was derived from the study in the same setting as the current study [7]. The post primary series GMC (95% CI) at 2 weeks after the second dose of the 2-dose homologous CoronaVac, 4 weeks apart, was 164.4 (133.55, 202.43); and after 2-dose homologous ChAdOx1, 10 weeks apart, was 278.5 (195.66, 396.33).
CI: confidence interval; IQR: interquartile range

